# Epidemic Overdispersion Strengthens the Effectiveness of Mobility Restrictions

**DOI:** 10.1101/2021.01.22.21250303

**Authors:** Gerrit Großmann, Michael Backenköhler, Verena Wolf

## Abstract

Human mobility is the fuel of global pandemics. In this simulation study, we analyze how mobility restrictions mitigate epidemic processes and how this mitigation is influenced by the epidemic’s degree of dispersion.

We find that (even imperfect) mobility restrictions are generally efficient in mitigating epidemic spreading. Notably, the effectiveness strongly depends on the dispersion of the offspring distribution associated with the epidemic. We also find that mobility restrictions are useful even when the pathogen is already prevalent in the whole population. However, also a delayed implementation is more efficient in the presence of overdispersion. Conclusively, this means that implementing *green zones* is easier for epidemics with overdispersed transmission dynamics (e.g., COVID-19). To study these relationships at an appropriate level of abstraction, we propose a **spatial branching process** model combining the flexibility of stochastic branching processes with an agent-based approach allowing a conceptualization of locality, saturation, and interaction structure.

## 1 Introduction

In 2020, the COVID-19 pandemic emerged and countries all over the world discussed which non-pharmaceutical interventions (NPIs) to implement to suppress or mitigate the spread of the novel SARS-CoV-2 pathogen [1]. Currently, in early 2021, a similar situation arises due to the uncertainty surrounding the appearance of the novel strains B117, 501yv2, and P1 [8]. Mobility restrictions are an important class of interventions, ranging from closures of international borders over restrictions to stay within a certain radius of one’s home to complete stay-at-home orders. The hope is that these measures constrain local outbreaks and that the virus reaches fewer susceptible host populations. Mobility restrictions could also help in creating so-called *green zones* within an infected population [5,6]. However, the effectiveness of NPIs depends on the epidemic’s properties.

*Overdispersion* is a property of an epidemic’s propagation. Specifically, each infected individual creates a certain number of *secondary infections* (aka *offspring*). This number depends on many factors and can be modeled using a discrete probability distribution called *offspring distribution*. The mean of the offspring distribution at time point *t* is the effective reproduction number *R*_*t*_. In the theory of stochastic branching processes, this probability distribution is the same for all agents and is independent of time *t* [4]. The offspring distribution can be characterized in terms of its *dispersion* (a measure of variance w.r.t. the mean). In the presence of high dispersion, many individuals produce zero or very few offspring while few individuals (so-called *supers-spreaders*) infect many others. Typically, the offspring number is generated as follows (i): sample an individual reproduction number *R*_*i*_ from a Gamma distribution with mean *R*_0_ and shape parameter *k* and (ii): sample the offspring number of agent *i* using a Poisson distribution with mean *R*_*i*_. Dispersion is typically quantified using the shape (aka dispersion) parameter *k* (smaller *k* implies higher dispersion) [4]. Combining a Gamma with a Poisson distribution results in a negative binomial distribution (NBD). For COVID-19 it has been reported that 80% of new infections can be traced back to only 15% of infected individuals relating to a *k* around 0.3 [7] (earlier work found *k* = 0.1 [2] and *k* = 0.25 [9]). Of particular importance for this work is the association of high dispersion with an increased die-out probability. Hence, it has been speculated that SARS-CoV-2 has to be introduced multiple times to a susceptible population in order to ignite an outbreak [3]. This property suggests a higher efficiency of mobility restrictions.

## 2 Spatial Branching Process

The default model type to study dispersion is a stochastic branching process [4] (BP). The state of a BP is a tree that grows over time. The children of a node correspond to the individuals infected by that node (i.e., its offspring). In each step, the number of offspring is sampled for each leaf. The epidemic is over when all leaves have zero offspring. An advantage of the BP model is the small number of parametric assumptions (all relevant aspects of the pathogen/environment are modeled with the offspring distribution). The BP model has two disadvantages for our purposes. There is no inherent form of locality in the model and no natural saturation (typically, an epidemic slows down when more agents become recovered/immune).

We account for this by placing the population in a Euclidean space. The number of offspring (offspring candidates to be more precisely) is still sampled from an offspring distribution but the spatial relationship influences which individuals are chosen. Moreover, an infection attempt is rejected if the new offspring candidate is already immune or infected. To model spatial movements more explicitly, we can randomly reposition agents. Note that the set of agents is fixed from the beginning (cf. Fig. 1 for an example).

**Fig. 1:**
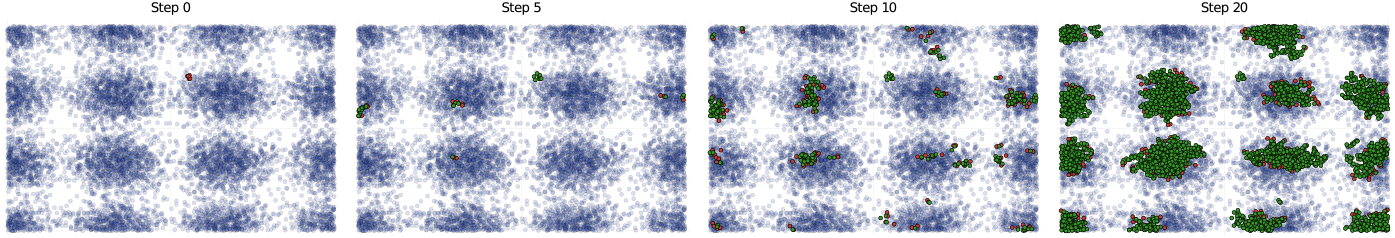
Example trajectory with Gaussian spatial kernel with *σ* = 0.008, two offspring-candidates, and *p*_*t*_= 0.1 (until 200 agents are recovered, then *p*_*t*_= 0.0).

### Model State

The global state is given by a set of agents *A*. Each *a*_*i*_ ∈ *A* is annotated with a position *x*_*i*_ ∈ [0, 1]^2^ and a local state *s*_*i*_ ∈ {*S, I, R*} (susceptible, infected, recovered). We initialize *x*_*i*_ according to a 2*d*-density *ν*.

### Model Dynamics

The global state changes randomly in discrete-time according to a discrete univariate offspring distribution *α*, a spatial interaction kernel *β*: ℝ_*≥*0_ → ℝ_*≥*0_, and a travel probability *p*_*t*_ ∈ [0, 1]. In each time step, for each infected agent *a*_*i*_:

1. Reposition *a*_*i*_ with probability *p*_*t*_ according to *ν*.
2. Generate offspring candidates, denoted *O*_*i*_ ⊂ *A* and infect all susceptible agents in *O*_*i*_.
3. Set *s*_*i*_ = *R*.

Regarding step 2: The offspring-count |*O*_*i*_| is sampled from *α*. Given |*O*_*i*_|, choose each *a*_*j*_ (*i* ≠ *j*) to be in *O*_*i*_ with a probability proportional to *β*(|| *x*_*i*_ − *x*_*j*_ ||).

## 3 Experiments

We use 10^4^ agents, *R*_0_ = 2.0, a Gaussian spatial kernel, and a mixture of 16 2*d*-Gaussians to generate the spatial positions (to mimic some population structure). Results are given in Fig 2. We compare a *fixed* offspring distribution where the (unsaturated) offspring count is always *R*_0_ = 2 with an NBD with varying *k*. Note that *k* =∞ leads to a Poisson offspring distribution as the variance of the Gamma distribution converges to zero. In the first experiment, we vary the variance, *σ*, of the spatial interaction kernel and set *p*_*t*_ = 0. Thereby, we can directly measure the influence of locality (smaller *σ* implies higher locality). In the second experiment we fix *σ* = 0.007 and vary *p*_*t*_. The value *σ* = 0.007 is such that the epidemic still dies out early with high probability for all offspring distributions with mean 2.0. This way, we measure locality by explicitly accounting for mobility. In the third experiment, we study delayed mobility restrictions. That is, we wait until 200 agents are infected (over varying *p*_*t*_), and then set *p*_*t*_ = 0.

**Fig. 2:**
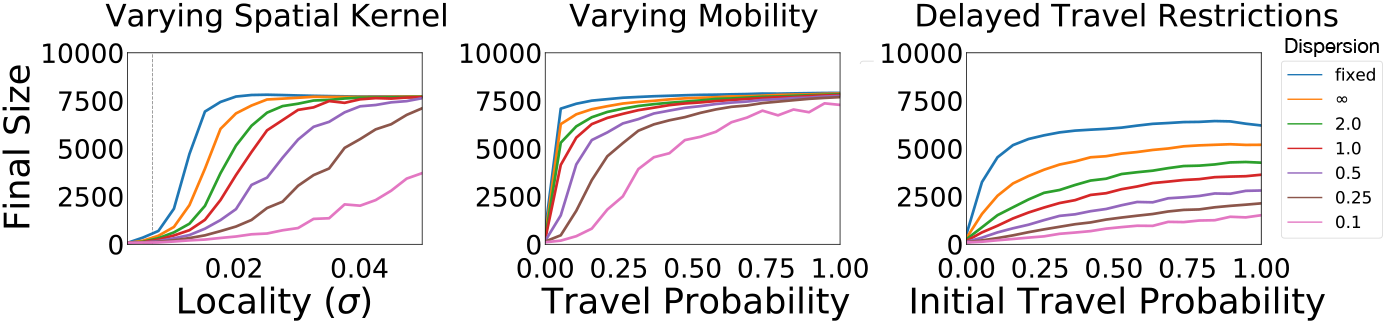
F.l.t.r.: Preliminary results for Exp. **1** to Exp. **3**. *x*-axis: Locality/mobility measure. *y*-axis: Final epidemic size/herd immunity threshold. Color indicates dispersion (smaller *k* implies higher dispersion).

The experiments consistently show that reducing mobility (in terms of *σ* or *p*_*t*_) has a stronger impact on the final epidemic the more dispersion is present in an epidemic’s transmission dynamics. This applies even if the mobility restriction is imperfect or implemented with some delay. Julia code is available^1^.

## 4 Conclusion

The relationship between dispersion, branching processes, and locality is under-explored in literature. We hope to spark interest in theoretical and practical work in this matter. For instance, it would be worthwhile to investigate if one can find optimal borders or levels on which mobility restrictions constitute the best trade-off between social costs and effectiveness. Understanding how to implement hierarchical mobility restrictions is also largely an open problem. Conclusively, this work can be seen as evidence that implementing a green zone strategy might be easier for epidemics that admit a higher dispersion.

## Data Availability

This is a theoretical study using only synthetically generated data. The procedures are described in detail in the manuscript.

https://github.com/gerritgr/SpatialBranchingProcess

## Acknowledgements

This work was partially supported by the DFG project MULTIMODE.

## Footnotes

1 github.com/gerritgr/SpatialBranchingProcess

